# Investigating shared genetic overlap of immune-mediated inflammatory diseases and cardiometabolic diseases

**DOI:** 10.64898/2026.06.09.26355246

**Authors:** Saleh Alduhayhi, Andrew P Morris, Sizheng Steven Zhao, John Bowes

## Abstract

**Background:** Immune-mediated inflammatory diseases (IMIDs) are associated with increased risk of cardiometabolic diseases. Investigating genetic overlap among these conditions can provide insights into their clinical management.

**Methods:** Genetic correlation was assessed using linkage disequilibrium score regression (LDSC). Then, a meta-analysis was conducted using Association Analysis Based on SubSETs (ASSET) to pinpoint independent single nucleotide polymorphisms (SNPs) shared across the diseases. Each independent SNP was then used to define a genomic window (+/-500KB) for colocalisation analysis and Local Analysis of [co]Variant Association (LAVA) to offer multiple layers of regional pleiotropic evidence. Over-representation analysis was then run to identify enriched biological pathways, which then were used for drug target analysis.

**Results:** The LDSC analysis showed a significant global genetic correlation for rheumatoid arthritis (RA) and cardiometabolic diseases including hypertension, coronary artery disease (CAD), heart failure (HF), stroke, atrial fibrillation (AF), and type two diabetes mellitus (T2DM) ranging from rg = 0.09 to 0.24. ASSET meta-analysis identified 164 independent SNPs shared across RA and the cardiometabolic diseases with *P* < 5×10-8 in the overall one-sided meta-analysis *P*-value, FDR<0.05 in both individual GWASs, and TRUE phenotype matrix. Colocalisation analysis revealed multiple loci with strong evidence (Posterior probabilities ≥ 80) of single causal SNPs between the trait pairs. LAVA analysis was then used as an additional layer of confirmation for the findings generated by ASSET and colocalisation and thus several loci were highlighted. Over-representation analysis showed significant enriched immune-related pathways across RA-hypertension, RA-CAD, RA-AF, and RA-T2DM trait pairs. Drug target analysis highlighted several drugs which could be further tested for their effectiveness in RA and its common comorbidities.

**Conclusion:** The findings revealed a shared genetic architecture and key immune-related biological pathways underlying RA and its associated cardiometabolic comorbidities. The identified genes and drugs provide opportunities for further therapeutic assessment which could improve clinical management strategies.

## Introduction

Immune-mediated inflammatory diseases (IMIDs) can be conceptualised as a group of heterogenous chronic conditions such as rheumatoid arthritis (RA), psoriatic arthritis (PsA), juvenile, and idiopathic arthritis (JIA), characterised by immune-driven inflammation.^1, 2^ The risk of developing cardiovascular diseases (CVDs) among IMID patients is observed.^2, 3, 4^ For example, the risk of developing CVDs is estimated to be 1.6 times greater in RA patients compared to the general population.^5^ Also, several observational studies have indicated that patients with RA are at increased risk of other comorbidities such as cardiometabolic diseases including coronary artery disease (CAD), type 2 diabetes mellitus (T2DM), and chronic kidney disease (CKD).^6^ This might be attributed to shared risk factors, chronic inflammation, sedentary lifestyle, and absence of appropriate anti-inflammatory drug and corticosteroid interventions.^7, 8, 9, 10, 11^

Prior studies^6,7, 8, 9, 10, 11^ have showed shared genetic components between IMIDs and cardiometabolic diseases. However, there has been no comprehensive approach to regionally and biologically investigate the shared genetics across IMIDs and their associated cardiometabolic comorbidities. Therefore, this study aims to examine genetic correlation of IMIDs and their cardiometabolic comorbidities, identify shared lead SNPs and genes, investigate shared enriched biological pathways, and prioritise shared candidate drugs.

## Method

### Study design

This research investigated the genetic overlap of IMIDs (RA, PSA, and JIA) and their associated cardiometabolic comorbidities including CKD, hypertension, CAD, heart failure (HF), stroke, atrial fibrillation (AF), and T2DM^12, 13, 14, 15, 16, 17, 18^, using genome wide association studies (GWAS) from European ancestry (Further details about the GWASs used are available in Supplementary Table 1). All analyses were run in R and the research methodology was a pairwise format organised into two main components conducted in multiple steps:

I. Linkage Disequilibrium Score Regression (LDSC)^19, 20^ for global heritability and genetic correlation
II. Association Analysis Based on SubSETs (ASSET) for cross-trait meta-analysis.^21^
III. Colocalisation analysis to assess whether RA and the associated comorbidities share single causal variants.^22^
IV. Local Analysis of [co]Variant Association (LAVA)^23^ for local genetic correlation
V. Multi-marker Analysis of GenoMic Annotation (MAGMA) for gene-based association analysis.^24^
VI. Over-representation enrichment analysis for exploring biological pathways.^25^
VII. Pathway pairing score for drug target analysis (Figure 1).^26^

**Figure 1.**
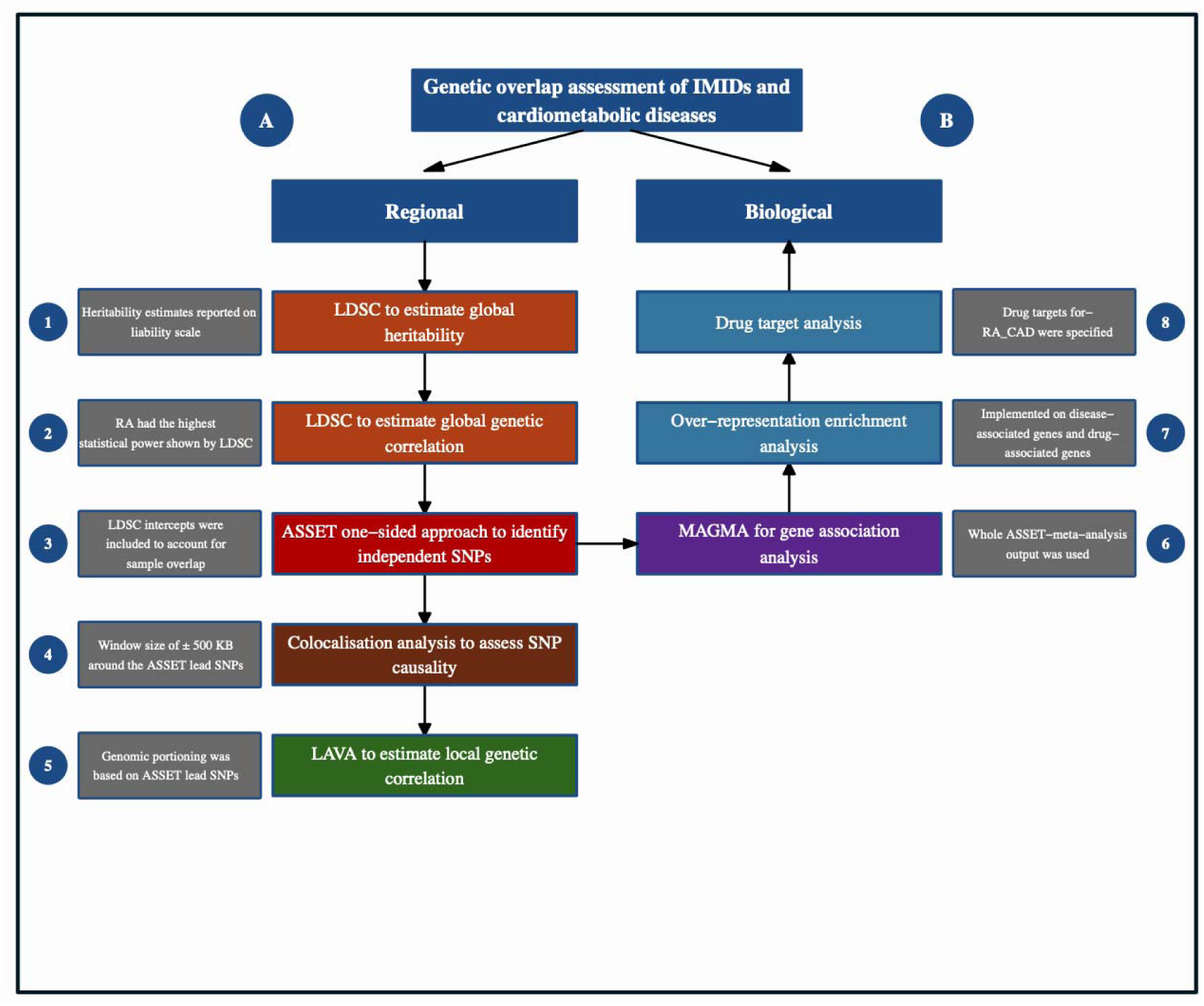
Illustration of the two main components-regional and biological. Trait pairs receiving consistent regional evidence across ASSET, colocalization, and LAVA were flagged as robust findings.

### Study population

Publicly available GWAS summary statistics across multiple traits were analysed. RA data were obtained from two corrected meta-analysed GWASs performed by METACARPA to account for unknown sample overlap.^27, 28, 29^ The first one was a multiomics GWAS conducted by Saevarsdottir et al. including 1,021,468 European individuals (31,313 cases and 995,377 controls)^30^. The second GWAS was performed by Ishigaki et al. where they analysed 97,173 European ancestry individuals (22,350 RA cases and 74,823 controls). RA cases were diagnosed based on the 1987 American College of Rheumatology (ACR) criteria, the 2010 ACR/European League Against Rheumatism criteria, or by a professional rheumatologist.^31^ PsA summary statistics were obtained from Soomro et al. GWAS incorporating 3,609 cases and 9,192 controls.^32^ JIA data were derived from a GWAS run by López-Isac et al consisting of 2,190 cases and 9,196 controls^33^ (Further details on the inflammatory diseases as well as the other cardiometabolic diseases are provided in Supplementary Table 1).

### Global heritability and genetic correlation

LDSC was used to estimate global heritability (*h*^2^) and genetic correlation (*r*_g_) between IMIDs such as RA, PA, and JIA and the cardiometabolic diseases. The entire distribution of p-values (not only significant ones) was analysed to capture all SNPs with small effects across the genome. LDSC regresses GWAS test statistics on LD scores to produce heritability estimates. This goes in line with genetic correlation assessment but with assessing LD scores and the test statistics of SNPs from different two GWAS summary data. Precomputed LD scores which were estimated from 1000 Genomes Phase 3 restricted to HapMap3 were used in the analysis https://utexas.app.box.com/s/vkd36n197m8klbaio3yzoxsee6sxo11v)^34^. LD score regression intercept distinguishes test statistic inflation not attributable to polygenic signal. Therefore, the intercept estimates can be used in various analyses to account for epidemiological biases such as sample overlap. An IMIDs with the highest statistical power was considered for the downstream analyses.^19, 20^

### Cross-trait-meta-analysis

A subset-based pleiotropic meta-analysis was performed using ASSET package in R to identify one-sided pleiotropic SNPs shared across RA and the cardiometabolic diseases. Generally, the one-sided version of this meta-analysis can detect SNPs with effects in consistent directions across different subsets of studies while in the two-sided approach associations can differ in direction. Overall, summary-statistics of heterogeneous GWASs were used to extract shared reference SNP cluster IDs (rsIDs), SNP regression betas and their standard errors. Given the disproportionate number of cases to controls, effective sample size was internally calculated. To account for sample overlap, a constructed KxK matrix estimated by LDSC supplied directly to ASSET. Multiple testing was automatically adjusted for using discrete local maxima (DLM), a method used to accurately estimate the probabilities of test statistics when searching across different subsets of studies. Therefore, the adjusted *P*-values can be interpreted as evidence of an overall association for the SNP across the subsets of selected studies. The returned combined effect sizes (betas) and their standard errors generated by the one-sided meta-analysis were computed using the standard fixed-effect meta-analysis across the selected studies.^21^

All one-sided SNPs were further clumped to ensure independent SNPs shared across RA and the cardiometabolic diseases (PLINK clumping, https://github.com/mrcieu/gwasglue#reference-datasets, parameters: --clump-p1 5e-8”, --clump-p2 1e-5, --clump-r2 0.1, --clump-kb 1000). The independent SNPs were visualised using UpSet plot generated with ‘UpSetR’ package.

SNPs were considered pleiotropic and significantly independent for two specific traits such as RA-T2DM, RA-CAD, or RA-hypertension etc … if they met the following criteria: 1. genome-wide significance (*P* < 5×10-8) in the overall one-sided meta-analysis *P*-value 2. FDR<0.05 in both individual GWASs and 3. inclusion of both traits in the optimal subset selected by ASSET (i.e., both traits had a “TRUE” entry in the phenotype matrix).

### Colocalisation analysis

Colocalisation analysis was performed using *coloc R package* to examine whether the associated SNPs for RA and the cardiometabolic diseases colocalise, with a particular focus on H4 (both traits share one causal variant). All genetic variants within ±500 kb of the ASSET independent SNPs were included in the analysis (potential parameters for both traits: beta, standard errors, sample size for proportion of cases, and rsIDs). Default prior probabilities were used (P1 = 1×10-4, P2 = 1×10-4, P12 = 1×10-5). Posterior probabilities ≥ 80 were considered as evidence of significant colocalisation.^22^

### Local genetic correlation analysis

LAVA was used to locally estimate genetic correlation of RA and the cardiometabolic diseases (input: PLINK format LD reference data, ASSET’s lead SNPs for locus definition, LDSC intercepts to account for sample overlap, and GWAS summary statistics). Briefly, LAVA uses GWAS effect estimates and LD structure from the 1000 Genomes Project reference panel to estimate the joint SNP effects which can then be used to estimate the local heritability from the local residual phenotypic variance. Local genetic correlation is calculated by forming a matrix of local genetic components from the joint effects on each trait, then estimating local genetic covariance which is then used to obtain the local genetic correlation.^23^

### Gene association analysis

MAGMA is a statistical tool that is based on a multiple regression model to properly adjust for LD between markers and to pinpoint multi-marker effects. It uses an F-test to calculate the gene *P*-value estimates in relation to traits of interest. Briefly, MAGMA transforms SNP data from summary statistics for a gene into a new set of principal component variables.

Then, it removes those with very small eigenvalues to use the remaining principal components as predictors of the trait in a linear regression model. MAGMA (*v1.10, custom*) was run using default parameters with a focus on European ancestry panel from the 1000 Genomes Project Phase 3 as the LD reference.^24^ Full pair-wise meta-analysis by ASSET was used in the MAGMA gene-level association analysis. To ensure the downstream analyses (over-representation and drug target analyses) were based only on genes with significant evidence of association with both traits, separate gene-based association analyses of individual traits were performed. Thus, a gene was considered significantly associated with a trait pair if it reached statistical significance at an FDR threshold of<0.05 in the gene analyses of ASSET meta-analysis and individual GWASs.

### Pathway enrichment analysis

Over-representation analysis was performed using “*clusterProfiler”* R package (Gene Ontology [GO] and Kyoto Encyclopedia of Genes and Genomes [KEGG]) for disease/drug-associated genes. Accounting for multiple testing was implemented using Benjamini–Hochberg (FDR<L0.05).^25^

### Drug target analysis

A pathway pairing score approach was used for this drug target analysis. Concisely, this approach retains the top 50 significant pathways of disease-associated genes as well as the drug-targeted genes (ranked by smallest adjusted P-value ^6^). These pathways were then partitioned into two tiers: Tier 1 consists of the top 20 terms and Tier 2 of the remaining pathways (21-50). A pathway which appears in Tier 1 of both lists (disease-associated pathways and drug-associated pathways) receives a full score of 1; a pathway appearing in Tier 1 of one list and Tier 2 of the other one receives a score of 0.5. The pathway pairing score was then developed by summing all full and half scores normalised by 20 for comparability. This gave an overall score ranging from 0 to 1. Higher pathway scores due to multiple overlapping pathways indicate greater biological similarity.^26^

In this study, pathway analyses were done for each trait pair. Any trait pair with no sufficient number of biological pathways was excluded. Disease-associated genes shared across trait pairs (RA-hypertension, RA-stroke, and RA-T2DM) were used to identify candidate drugs (i.e. the candidate drugs should target the disease-associated genes), using drug-gene databases such as DrugBank^35^, DrugCentral^36^, DGIdb^37^, and PharmGKB^38^. The analysis was restricted to drugs that have a relationship with RA and the cardiometabolic diseases. Then, the four drug-gene databases were again queried to retrieve all genes targeted by these final candidate drugs. ClusterProfiler was then used to perform a biological pathway analysis on the drug-associated genes. Finally, pairing scores were computed between disease-associated pathways (trait pairs) and drug-associated pathways (the threshold set for a candidate drug to be significant was a score of ≥ 0.5).^39^

## Results

### Genetic correlation

LDSC analysis showed that all analysed IMIDs (RA, PA, and JIA) and cardiometabolic diseases including CKD, hypertension, CAD, HF, stroke, AF and T2DM captured a detectable polygenic signal to reliably run the downstream analyses. (Table 1).

**Table 1.**
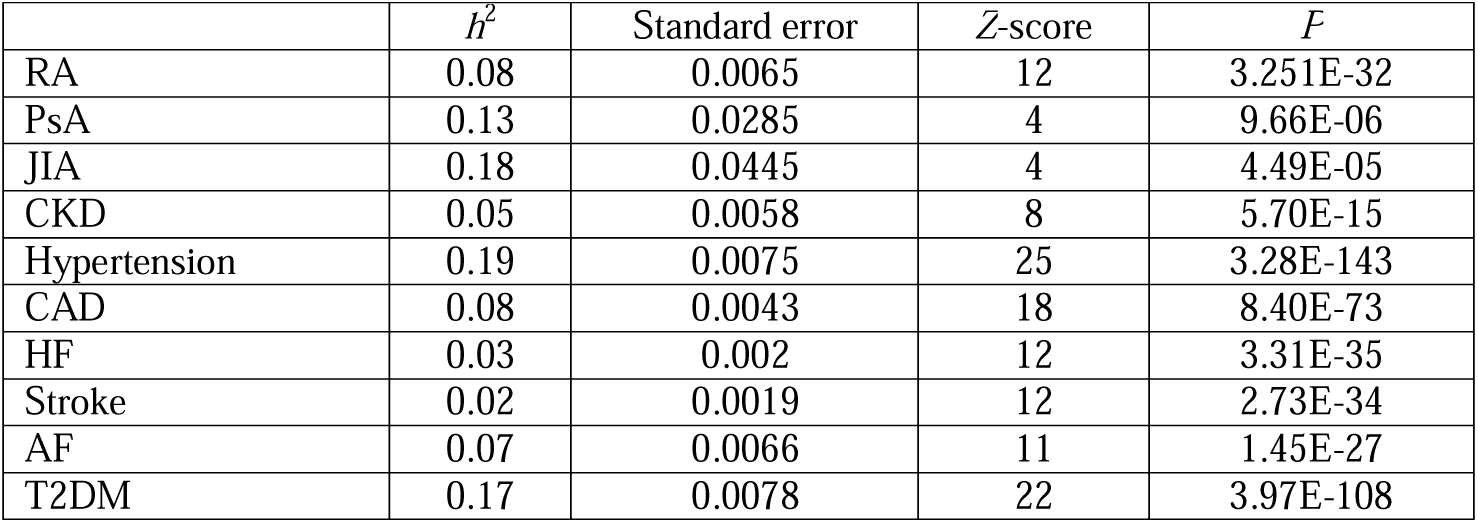
Heritability estimates (liability scale)

| | $h^2$ | Standard error | Z-score | *P* |
| --- | --- | --- | --- | --- |
| RA | 0.08 | 0.0065 | 12 | 3.251E-32 |
| PsA | 0.13 | 0.0285 | 4 | 9.66E-06 |
| JIA | 0.18 | 0.0445 | 4 | 4.49E-05 |
| CKD | 0.05 | 0.0058 | 8 | 5.70E-15 |
| Hypertension | 0.19 | 0.0075 | 25 | 3.28E-143 |
| CAD | 0.08 | 0.0043 | 18 | 8.40E-73 |
| HF | 0.03 | 0.002 | 12 | 3.31E-35 |
| Stroke | 0.02 | 0.0019 | 12 | 2.73E-34 |
| AF | 0.07 | 0.0066 | 11 | 1.45E-27 |
| T2DM | 0.17 | 0.0078 | 22 | 3.97E-108 |

Significant genetic correlations (*P* < 0.05) were observed among RA, PsA, and JIA. RA was the only inflammatory disease exhibiting a significant positive genetic correlation with six cardiometabolic diseases including hypertension, CAD, HF, stroke, AF, and T2DM. By contrast, JIA demonstrated significant genetic correlation with hypertension, CAD, HF, and T2DM only. Compared to the other inflammatory diseases, PsA showed a minimal significant genetic correlation with CAD but a higher genetic estimate with T2DM. None of the inflammatory diseases exhibited genetic correlations with CKD. Among the cardiometabolic diseases, all traits showed highly significant genetic correlations with one another, with CAD and HF demonstrating the highest genetic overlap, whereas CKD exhibiting less genetic correlations; CKD was only associated with hypertension, CAD, and HF. RA was the only inflammatory disease selected for the subsequent analyses due to its higher statistical power (larger sample size and Z score) and it has genetic correlation with the majority of cardiometabolic diseases except CKD, compared to other inflammatory diseases (Figure 2).

**Figure 2.**
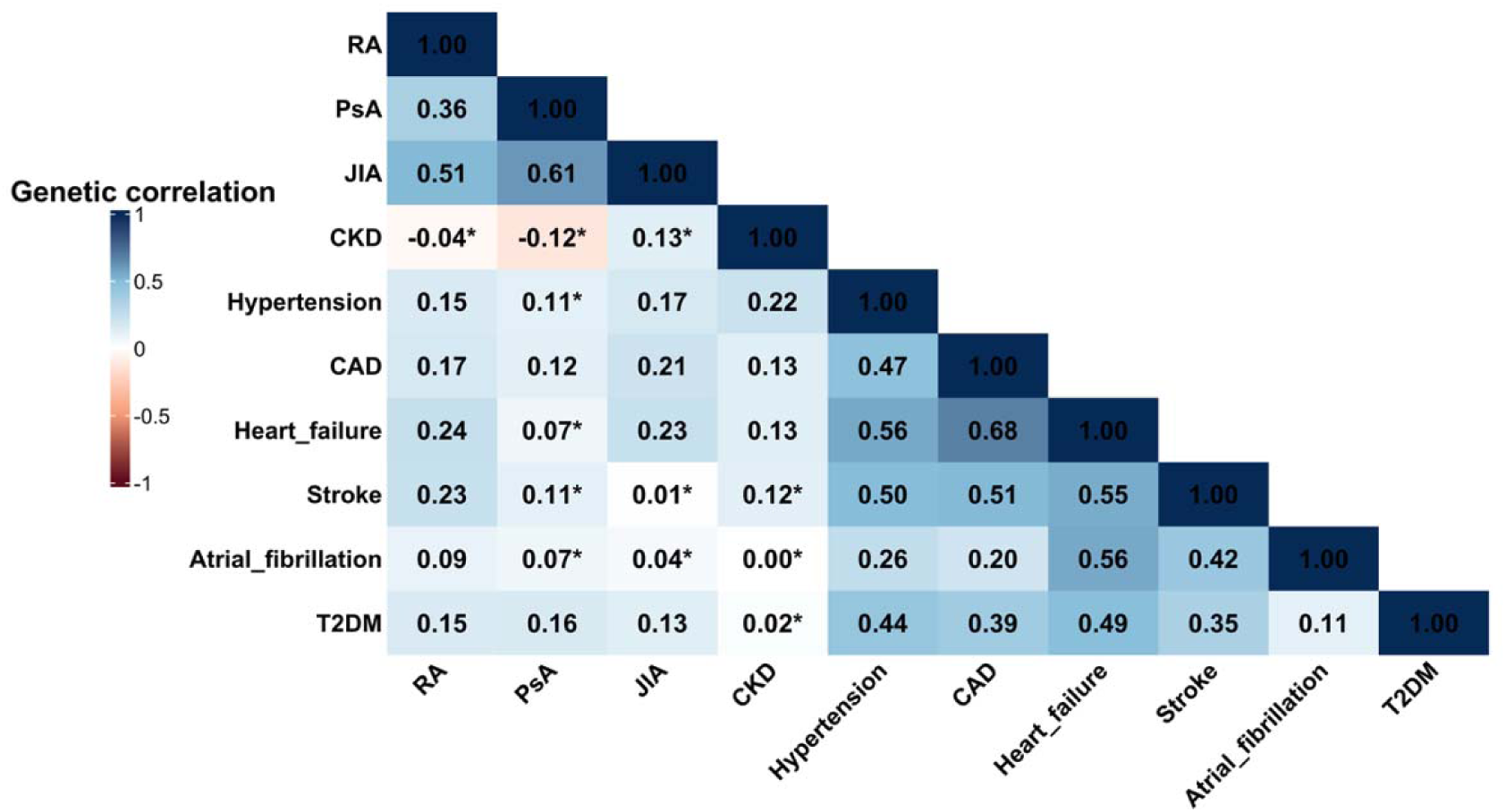
Global genetic correlation estimates of IMIDs and cardiometabolic diseases. Asterisks indicate that the estimate did not reach statistical significance *P* < 0.05. Light/dark orange means the estimate is on the opposite direction (inverse genetic correlation).

### ASSET cross-trait meta-analysis, colocalisation, and LAVA

Evidence of pleiotropic loci shared a cross RA and cardiometabolic diseases was established using complementary statistical methods. The pairwise cross-trait meta-analysis identified 164 independent significant SNPs shared across RA and all cardiometabolic diseases except for CKD: 42 lead SNPs linked to RA-hypertension, 42 for RA-CAD, 1 for RA-HF, 7 for RA-stroke, 19 for RA-AF, and 53 for RA-T2DM (Figure 3).

**Figure 3.**
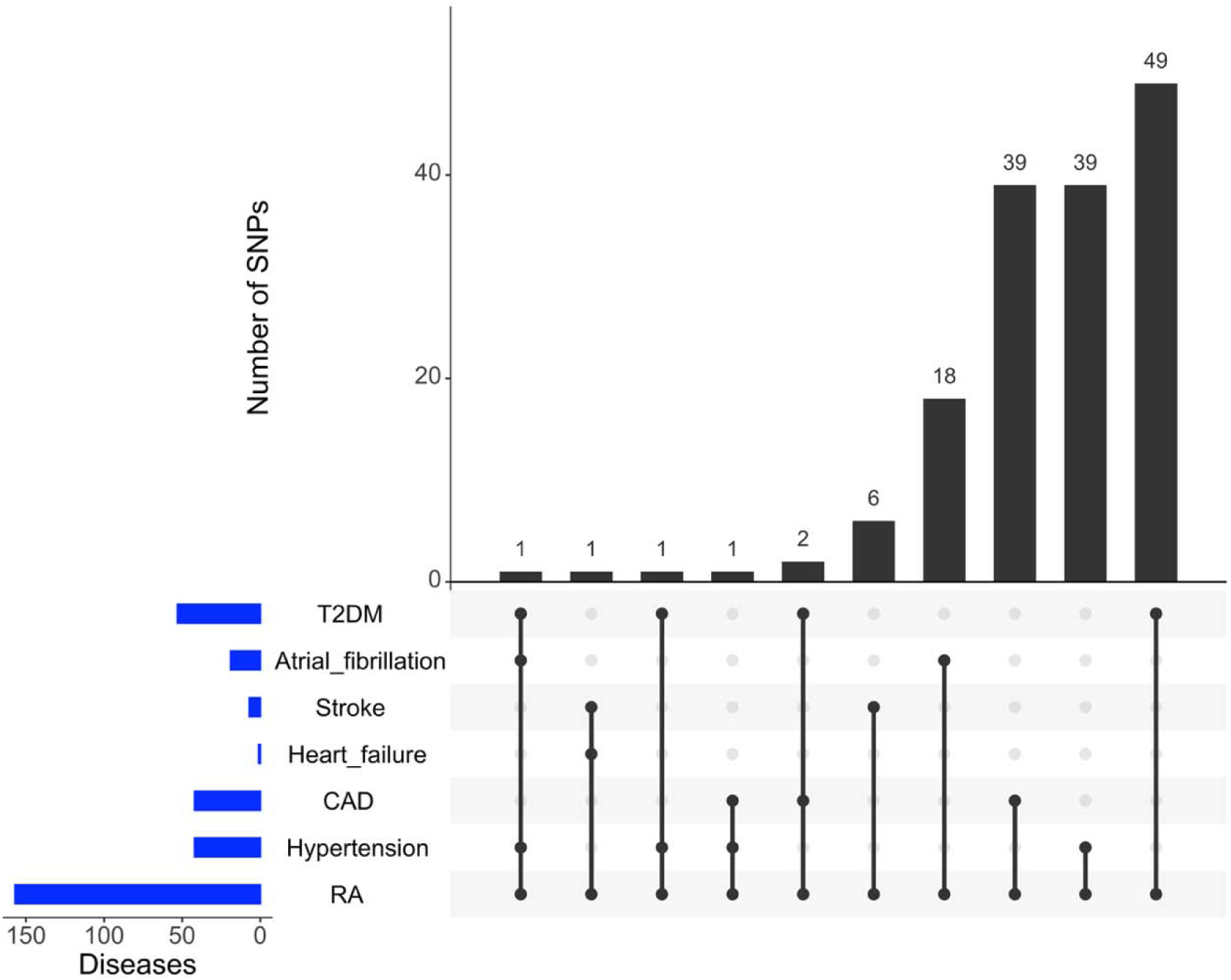
ASSET meta-analysis of RA and the cardiometabolic diseases. Significant loci were defined at ASSET_meta_*P*-value < 5×10-8, FDR<0.05 in both individual GWASs, and both traits were marked as “TRUE” entry in the ASSET phenotype matrix.

Supplying the ± 500 KB window around the lead SNPs to colocalisation and LAVA analyses identified significant loci. Significant loci were defined as genomic regions that met all three criteria: (1) they originated from the ASSET independent SNPs, (2) they demonstrated strong evidence of colocalisation H4 ≥ 80%, (3) they showed significant positive local genetic correlation by LAVA (*P* = < 0.05). Therefore, four loci were identified for RA-Hypertension (chr11:9770910, chr12:112023001, chr16:28922149, and chr12:111361298). Seven loci for RA_CAD (chr12:111910219, chr12:111281636, chr12:111883779, chr12:111631733, chr12:113168368, chr12:112941571, and chr1:114069639). A single locus at chr12: 111973358 with its interval of 111723358-112223358 had shown a pleiotropic implication in both RA-HF and RA-stroke while four distinct loci were detected for RA-T2DM (chr13:31042452, chr18:2114234, chr11:121983370, and chr16:28917644, Supplementary Table 2).

### Overlapping genes and pathways

After applying the predefined filtering criteria, the gene-based-association analyses suggested 422 genes shared pleiotropically across RA-hypertension, 320 genes identified in RA-CAD, 3 genes associated with RA-HF, 29 genes for RA-stroke, 123 genes for RA-AF, and 318 genes linked to RA-T2DM (Figure 4 - Supplementary Table 3-8).

**Figure 4.**
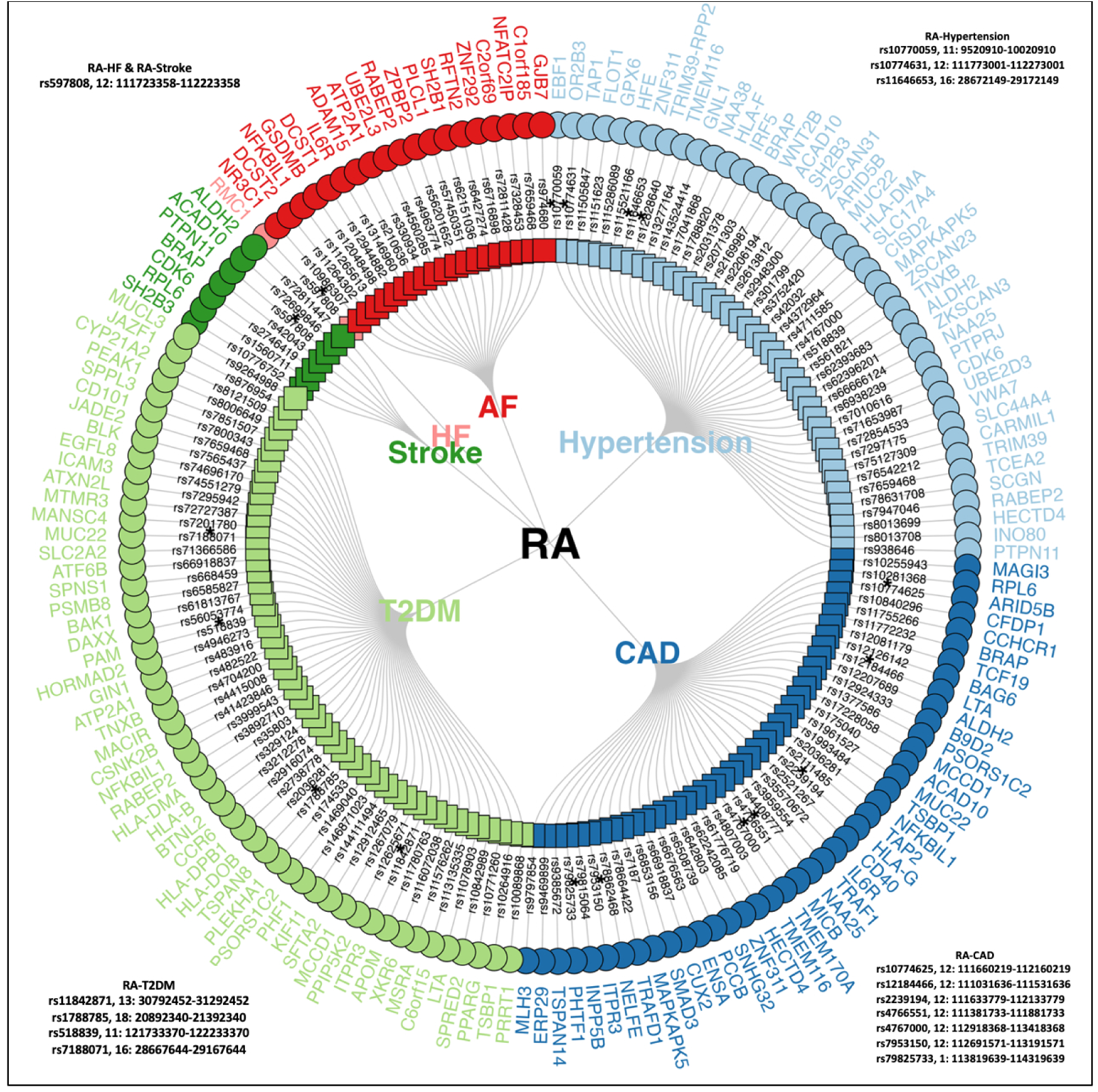
Summary of the most significant results by ASSET, colocalisation, LAVA, and MAGMA. Genetic variants in the first layer are lead SNPs shared across RA and the cardiometabolic diseases. An asterisk marks loci with strong multiple evidence by ASSET, colocalosation, and LAVA (small boxes around the figure summarise the significant loci supported by the three methods). Outer-layer indicates the top prioritised genes per trait-pair selected by the smallest *P*-values (FDR).

The over-representation analyses across RA-hypertension, RA-CAD, RA-AF, and RA-T2DM confirmed enriched pathways related to immune system (Figure 5). The gene sets for RA and the remaining cardiometabolic traits showed inadequate coverage to support strong pathway-level results.

**Figure 5.**
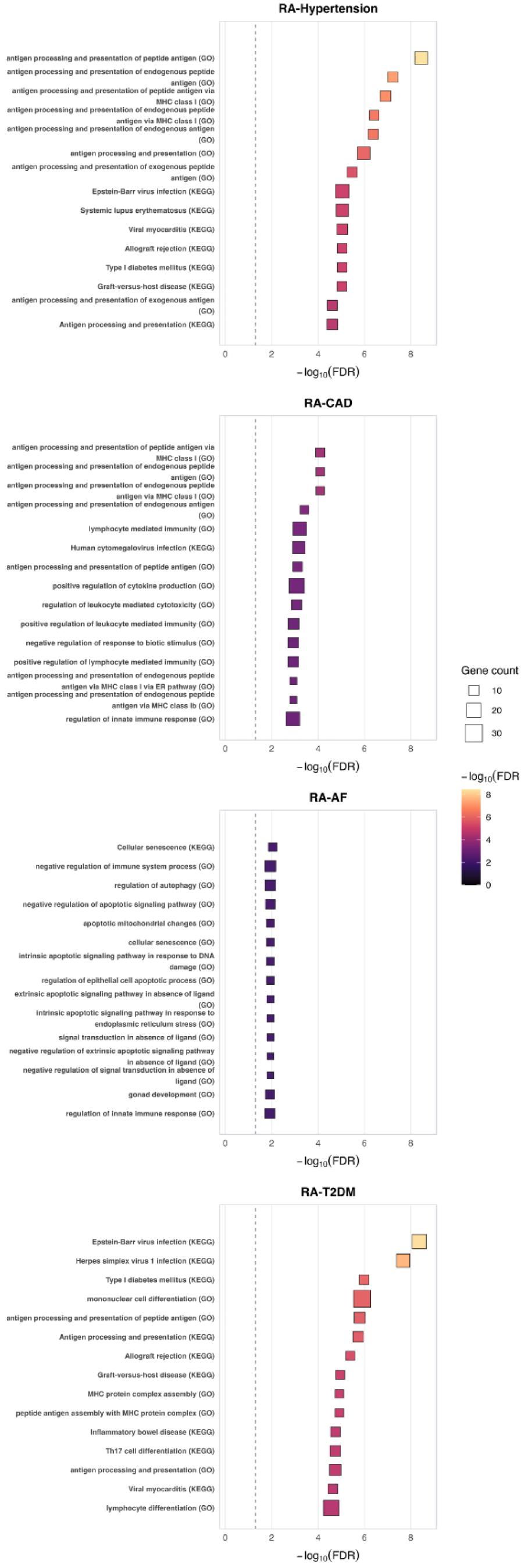
Immune pathways enriched across RA-hypertension, RA-stroke, and RA-T2DM with their associated gene count and FDR-adjusted *P*-values.

### Drug target analysis

790 disease genes shared across RA, hypertension, CAD, AF, or T2DM were identified. The remaining traits were excluded as their over-representation analyses did not produce sufficient biological pathways. The genes were then used to identify drugs relevant to RA and the associated comorbidities from the drug-gene databases. This has led to many candidate drugs, but the analysis was restricted to RA-cardiometabolic disease relevant drugs and thus, 3 candidate drugs were retained. Tocilizumab which belongs to Disease-Modifying Antirheumatic Drugs (DMARDs) was the one with highest pairing score which suggests a degree of overlap between its biological pathways and the enriched biological pathways of the RA_CAD associated genes (Figure 6)

**Figure 6.**
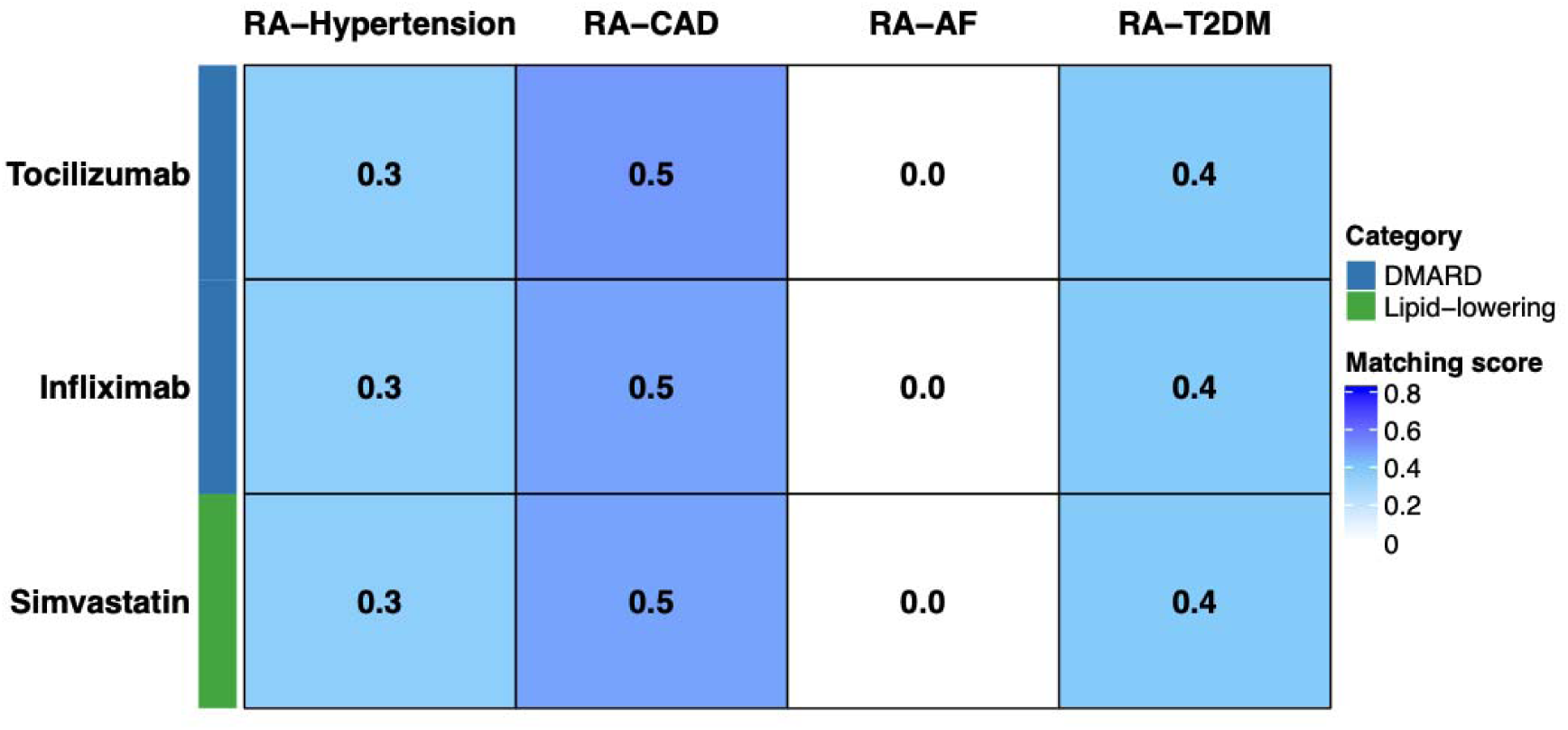
Demonstration of each drug’s pairing score with each trait pair. A score of ≥ 0.5 considered as potential drugs for further investigation.

## Discussion

This study provides a comprehensive, multi-layered investigation of the shared genetic architecture between IMIDs and cardiometabolic diseases, with RA exhibiting robust genetic overlap across multiple cardiometabolic outcomes. Using complementary global, regional, and biological approaches, significant genome-wide genetic correlations between RA and six cardiometabolic diseases were identified, alongside 164 independent pleiotropic loci supported by cross-trait meta-analysis, colocalisation, and local genetic correlation analyses. These shared loci converged on immune-related biological pathways and highlighted clinically relevant genes and drug targets, reinforcing inflammation-driven mechanisms as a key link between RA and its cardiometabolic comorbidities.

SH2B adaptor protein 3 (SH2B3) gene was found associated with RA-stroke pathogenesis. SH2B3 belongs to SH2B family consisting of another two adaptor proteins –SH2B1 and SH2B2. This gene is well known for signal regulation and haematopoiesis and immunity. Negative mutations in this gene can lead to myeloproliferative disorders and inflammatory diseases such as RA and stroke in mice.^40^ ^41, 42^ interleukin 6 receptor (IL6R) is a gene found implicated in RA-CAD which plays an important role in immune and inflammatory responses. When dysregulated, the persistent inflammation can be involved in the pathogenesis of RA and CAD. Therefore, IL6R represents a therapeutic target to manage and control inflammation. ^43, 44, 45^ The pathway-pairing score flagged several drug candidates (tocilizumab, infliximab, and simvastatin) for RA-CAD based on their biological similarity.

Tocilizumab is one of the biologic DMARDs that acts as an anti-interleukin-6 receptor inhibitor.^46^ It is commonly prescribed to RA patients and can be safely combined with conventional synthetic disease-modifying antirheumatic drugs (csDMARDs) such as methotrexate.^47, 48^ While Tocilizumab in a 16-week noninterventional prospective study of patients with RA led to significant improvement in endothelial function, it increased total cholesterol and low-density lipoprotein levels, which could indicate long-term cardiovascular risk uncertainty.^49^

Another biologic DMARDs is infliximab (anti-tumour necrosis factor alpha inhibitor) used to treat RA.^50^ Evidence on infliximab’s impact on CAD risk in RA patients is inconsistent and needs further validation especially that they are based on CAD surrogate markers such as pulse wave velocity, carotid intima-media thickness, and plaque. A post hoc analysis of longitudinal data obtained from a randomised placebo-controlled study aimed to assess the effects of infliximab treatment on vascular stiffness such as pulse wave velocity, carotid intima media thickness and carotid artery plaque in RA patients. The main findings showed that 56 weeks of infliximab therapy in RA patients significantly improved arterial stiffness (pulse wave velocity) but did not alter carotid intima media thickness and carotid artery plaque suggesting partial benefits for RA patients.^51^ In another observational study done by Micco et al. on 14 RA patients. They noticed that RA patients treated with infliximab had increased intima-media thickness and atherosclerotic plaques compared to those who were not on infliximab.^52^

Simvastatin is 3-Hydroxy-3-Methylglutaryl-Coenzyme A (HMG-CoA) reductase inhibitor commonly used to lower low-density lipoprotein cholesterol, triglycerides and apolipoprotein B while modestly increase high-density lipoprotein cholesterol.^53^ It can be prescribed to RA patients to indirectly minimise the risk of CAD (via total and LDL cholesterol level reduction). Generally, statins have pleiotropic anti-inflammatory and immunomodulatory effects that extend beyond lipid lowering. In a prospective cohort study including 100 RA patients who were given simvastatin combined with DMARD therapy over six months.

Patients receiving this drug showed slight improvement in RA disease activity as well as considerable reduction in total cholesterol and LDL-cholesterol. However, low dose of simvastatin may cause adjunctive anti-inflammatory effect in RA patients but insufficient to replace the use of DMARD. Its primary benefit remains in cardiovascular disease risk reduction among RA patients.^54^

This study has many strengths. Large-scale summary statistics were adopted for the analyses and quality control measures were further implemented to maximise accuracy. The genetic overlap was investigated through a multi-staged analytic framework which offered a thorough characterisation of regional-biological exploration of RA and its associated common comorbidities. The biological findings were produced through a clear clinical translational path (gene > pathway > drug target evaluation). The pathway-pairing score assessed how well candidate drugs match shared RA-CAD biology at the biological pathway level. For instance, the pathway alignment observed for Simvastatin was driven by overall lipid reduction in RA patients when combined with other DMARDs, indicating potential relevance for RA patients with elevated lipid levels without implying disease modifying effects.

A key limitation of the pathway pairing score is that while it quantifies biological similarity between drug-disease-related pathways, it does not specify the direction of effect (i.e., it does not tell if a candidate drug increases or decreases the risk of a disease, literature can help add insights into the direction). Furthermore, although the drugs that met the predefined threshold demonstrated strong biological overlap, this should not be interpreted as evidence that they are ready for clinal use. Instead, the analysis highlighted the candidates that were wroth further observational and experimental validation as well as clinical evaluation.

It is important to note that drugs that did not reach the predefined pathway-pairing score threshold (≥ 0.5) should not be interpreted as invalid or clinically inappropriate; a low score in this study for instance reflects limited pathway overlap with the specific gene sets used, not a lack of therapeutic value.

Furthermore, pathway boundaries or the set of genes are frequently arbitrary and vary between databases; analysing multiple sources can enhance result accuracy. The majority of enrichment analysis methods assume statistical independence of genes and pathways, which is not always achievable.^55^

Another disadvantage is that results cannot be generalised to other ancestries as the focus was on Europeans only. There is a potential sample overlap across the analysed GWASs which might inflate test statistics. However, sample overlap within RA GWASs was corrected using METACARPA while cross-trait LDSC intercepts showed little evidence of sample overlap biasing the produced RA-cardiometabolic estimates although these intercepts may also capture other residual biases such population stratification and cryptic relatedness^19, 20^ (Supplementary Table 9).

## Conclusion

The study integrated a comprehensive approach to explore the genetic overlap of IMIDs and their associated comorbidities. The analyses highlighted a significant regional and biological association between RA and its comorbidities. Pathway-pairing score analysis showed biological similarity between the disease-associated genes and the drug-associated ones.

Further experimental studies are needed to evaluate the identified drug effectiveness on RA-associated comorbidities.

## Ethical approval

This study was based on secondary analyses of publicly available GWAS summary statistics. All original GWASs generating these datasets were implemented in accordance with agreed ethical standards and each received approval from relevant institutional review boards or ethics committees. The present study did not involve any new data collection, direct participant contact, nor access to individual-level data and therefore, no new ethical approval was required.

## Data availability

The GWAS summary statistics analysed in this study are publicly available from their original sources. Details are provided in Supplementary Table 1. For RA data, the following links offer the datasets: Saevarsdottir S. et al. RA and its subtypes (https://www.decode.com/summarydata/)

Ishigaki RA (https://www.ebi.ac.uk/gwas/studies/GCST90132223),

PsA https://www.ebi.ac.uk/gwas/publications/35507331

JIA https://www.ebi.ac.uk/gwas/publications/33106285

CKD https://ckdgen.imbi.uni-freiburg.de/datasets/Wuttke_2019

Hypertension https://www.ebi.ac.uk/gwas/studies/GCST007610

CAD https://www.ebi.ac.uk/gwas/studies/GCST90132314

HF https://cvd.hugeamp.org/dinspector.html?dataset=GWAS_HERMES_eu&phenotype=HF

Stroke https://www.ebi.ac.uk/gwas/studies/GCST90104539

AF https://www.ebi.ac.uk/gwas/studies/GCST006414

T2DM https://t2d.hugeamp.org/dinspector.html?dataset=Mahajan2022_T2D_EU&phenotype=T2D

